# Efficacy of cardioneurablation for vasodepressor vasovagal syncope

**DOI:** 10.1101/2023.09.15.23295646

**Authors:** Zhenhuan Chen, Ying Li, Yuan Liu, Guowang Shen, Ganwei Xiong, Bo Wu, Yanfeng Liu, Xiantao Huang, Hongyan Li, Haiwen Zhou, Zhicheng Xu, Gulao Zhang, Yu Tao, Fanzhi Zhang, Hengli Lai

**Author notes:** Correspondence to: Hengli Lai, Email address, Department of Cardiology, Jiangxi Provincial People’s Hospital, The First Affiliated Hospital of Nanchang Medical College, Aiguo Road, Donghu District, Nanchang, Jiangxi 330000, China. Contribute equally in this study (Zhenhuan Chen, Ying Li).

## Abstract

**Objective:** Cardioneuroablation (CNA) is effective for cardiac inhibitory and mixed vasovagal syncope (VVS) but not vasodepressor VVS. This study aimed to assess the therapeutic benefits of CNA in vasodepressor VVS.

**Methods:** VVS patients hospitalized in the Department of Cardiology of Jiangxi Provincial People’s Hospital were retrospectively reviewed. Holter monitoring was performed before, during, and 3 months after CNA. Changes in heart rate and Wenckebach point of atrioventricular conduction before and after ablation were compared.

**Results:** Thirty-five patients were enrolled, 18 men and 17 women. Mean age was 47.48 ± 16.49 years. Median duration of syncope was 24.0 months (range, 2.5–66.0). Median number of syncope episodes before treatment was two (range, 2–4). The time domain indexes of heart rate variability, mean heart rate, maximum heart rate, and minimum heart rate were significantly higher 3 months after CNA. Mean follow-up was 11 ± 4.67 months. Recurrent syncope occurred in two patients with vasodepressor VVS, one of them with presyncope symptoms in vasodepressor type; and one patient occurred with mixed VVS, without presyncope symptoms. No serious complications occurred. CNA is safe and effective in the treatment of vasodepressor VVS.

**Conclusions:** CNA is effective for treating vasodepressor VVS. Our study provides a theoretical basis for individualization of treatment in patients with vasodepressor VVS.

## INTRODUCTION

Vasovagal syncope (VVS) is the most common type of syncope. Most patients who experience VVS also experience prodromal symptoms such as nausea, dizziness, fatigue, sweating, and/or amaurosis. The syncope occurs because of impaired autonomic regulation of the heart and consequent brain hypoperfusion[1]. The pathogenesis of VVS is complex and has not been fully elucidated. Several mechanisms may be involved, including the Bezold-Jarisch reflex, autonomic nervous dysfunction, neurohumoral factors, fluid-mediated vasodilatation, decreased baroreceptor sensitivity, heredity, and genetic factors[2]. The head-up tilt table test is usually used to diagnose and classify VVS based on heart rate and blood pressure during syncope. VVS is divided into three types, cardioinhibitory, vasodepressor, and mixed. Although generally benign, recurrent VVS can affect quality of life and result in patient injury[3]. Therefore, the main goal of treatment is prevention of recurrent events. Reported recurrence rates are as high as 61%[4–7].

Non-pharmacological treatments such as maintenance of adequate fluid and salt intake, avoidance of syncope triggers, and tilt training have no significant effect in many patients. Cardiac postganglionic parasympathetic neurons, which are mainly located in the epicardial fat pad, regulate cardiac rhythm and conduction[4, 8]. In preliminary studies, cardioneuroablation (CNA) was effective in treating VVS and achieved good long-term outcomes[9, 10]. However, its effectiveness varies among patients with different types of VVS. This study aimed to evaluate the safety and effectiveness of CNA for VVS using heart rate variability (HRV) indexes and recurrent syncope as outcome measures. We also compared the effectiveness of CNA between patients with vasodepressor and mixed types of VVS.

## MATERIALS AND METHODS

We retrospectively reviewed consecutive patients with VVS who were hospitalized in Jiangxi Provincial People’s Hospital from July 2020 to July 2022 and underwent CNA. Head-up tilt table testing was positive in all. VVS was diagnosed using the 2018 European Society of Cardiology Guidelines. In our institution, patients with recurrent syncope that seriously affects quality of life or causes injury who do not respond to conventional treatment are considered for CNA. Patients diagnosed with causes of syncope other than VVS were excluded; all underwent examinations to exclude structural heart disease, nervous system disease, and other causes. We also excluded patients with atrial thrombus on transesophageal echocardiography. All patients provided written informed consent.

### Head-up tilt table testing

The head-up tilt table test was considered positive in patients who experienced syncope, near syncope, or prodromal symptoms such as pallor, sweating, palpitations, nausea and vomiting, dizziness, or dyspnea in conjunction with a decrease in blood pressure and/or heart rate. The positive reactions were classified into the following types according to changes in blood pressure and heart rate:

1. type 1 (mixed type)—heart rate decreased during syncope but was ≥40 beats/minute or <40 beats/minute for less than 10 seconds, with or without cardiac arrest <3 seconds; blood pressure also decreased
2. type 2a (cardioinhibitory type)—heart rate <40 beats/minute for more than 10 seconds, no cardiac arrest; blood pressure decreased before heart rate
3. type 2b (cardiac depression type)—cardiac arrest >3 seconds; heart rate and blood pressure decreased at the same time or heart rate decreased before blood pressure did
4. type 3 (vasodepressor type)—systolic blood pressure <80 mm Hg or mean blood pressure decrease >30 mm Hg; heart rate decrease not more than 10% during syncope

Before the test, electrocardiographic monitoring and venous access were established. After remaining supine for 10 minutes, the patient’s heart rate and blood pressure were recorded; then, the bed was tilted 70° to begin the test phase.

### Mapping of the ganglionated plexuses (GPs)

There are five autonomic GPs in the left atrium. The left superior GP (LSGP) is located at the anterolateral junction of the left superior pulmonary vein and left atrium. The left inferior GP (LIGP) is located in the posterior inferior region of the junction between the left inferior pulmonary vein and the left atrium. The posteromedial left GP (PMLGP) is in the posterior atrial septum, between the left atrial posterior wall, inferior vena cava, and coronary sinus opening. The right anterior GP (RAGP) is in the anterior region of the junction between the right superior pulmonary vein and the left atrium. The right inferior GP (RIGP) is in the anterior region of the junction between the right inferior pulmonary vein and the left atrium. After anatomical localization, high-frequency stimulation (HFS) was used to determine the location of the left atrial vagal ganglion. When performing HFS near the mitral annulus, attention was paid to prevent malignant arrhythmias such as ventricular tachycardia or ventricular fibrillation. Vagal reflexes during HFS manifested as transient ventricular arrest, atrioventricular block, or 50% prolongation of the mean RR interval. Sites with positive vagal responses were recorded as GP targets.

### CNA

To perform CNA, the patient was positioned flat on the operating table and the skin of the head, neck, chest, and both inguinal regions was sterilized and draped. After injection of 2% lidocaine as a local anesthetic, the left and right femoral veins were puncturing using the Seldinger technique. The right ventricular and coronary electrodes were placed to induce an intracavitary electrocardiogram. Then, an 8.5 F SL1 SWARTZ sheath (Abbott Laboratories, Chicago, IL, USA) was inserted into the right atrium through the right femoral vein. Interatrial septal puncture was performed under fluoroscopy. After successful puncture, the patient was heparinized to maintain an activated clotting time between 200 and 300 ms. Then, the sheath was advanced into the left atrium and a three-dimensional anatomical model of the left atrium was constructed using the ablation electrodes under the guidance of the CARTO3 system (Biosense Webster, Irvine, CA, USA). The location of common cardiac ganglia were marked and HFS performed at each location. When a vagal response and hypotension were observed within a few seconds after ablation, the appropriate ablation site was determined. Power and temperature were set at 35-45W and 43°C, respectively. The ablation endpoint was no vagal response induced by repeated HFS.

### Statistical analysis

Statistical analyses were performed using SPSS software version 17.0 (IBM Corp., Armonk, NY, USA). Continuous data are expressed as means with standard deviation and were compared using one-way analysis of variance or the paired sample t test as appropriate. Categorical variables are expressed as numbers with percentage and were compared using the chi-square test. P <0.05 was considered significant.

## RESULTS

### Patient characteristics

Of 35 patients in all type of VVS, 13 were vasodepressor type, including seven men and six women. Mean age all type VVS patient was 47.48 ± 16.49 years. Median duration of syncope was 24.0 months (range, 2.5–66.0). Median number of syncope episodes before treatment was two (range, 2–4). Patient characteristics are shown in Table 1.

**Table 1.** Patient characteristics.

| Variables of interest | result |
| --- | --- |
| Age (year) | 47.48±16.49 |
| Male | 7/13 |
| History of syncope (year) | 4.14±4.45 |
| The number of preoperative syncope | 2.93±1.83 |
| Basal systolic blood pressure(mmHg) | 126.46±15.22 |
| Basal diastolic blood pressure(mmHg) | 76.46±12.34 |
| Basic heart rate | 71.19±10.40 |
| Results of the head-up tilt test |  |
| Positive Systolic Blood Pressure (mmHg) | 79.00±15.19 |
| Positive diastolic Blood Pressure (mmHg) | 44.46±11.43 |
| Heart rate during syncopal episode | 62.62±23.59 |
| Results of echocardiography |  |
| Left atrial anteroposterior diameter (mm) | 30.47±4.42 |
| Left ventricular end-diastolic diameter (mm) | 44.26±4.35 |
| Left ventricular ejection fraction (%) | 62.00±0.04 |

### Cardiac radiofrequency ablation

The ablation electrode was used to perform HFS at five common GP sites in the left atrium. Figures 1 and 2 show the GP distribution in the three-dimensional reconstruction model of two patients. Figure 3 shows a schematic diagram of one patient during ablation.

**Figure 1.**
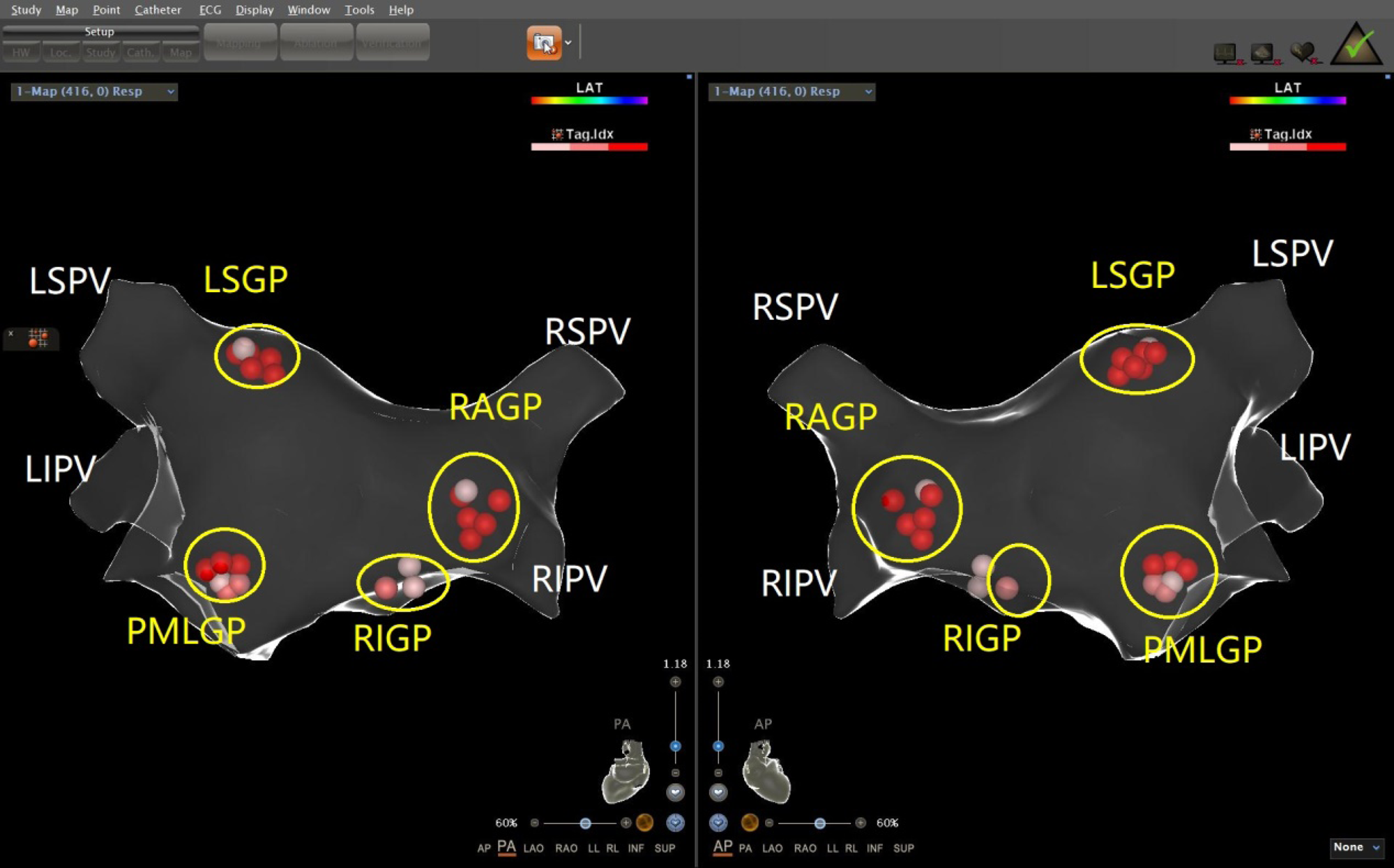
Schematic representation of the distribution of the autonomic ganglionated plexus in the left atrium of a patient with mixed VVS

**Figure 2.**
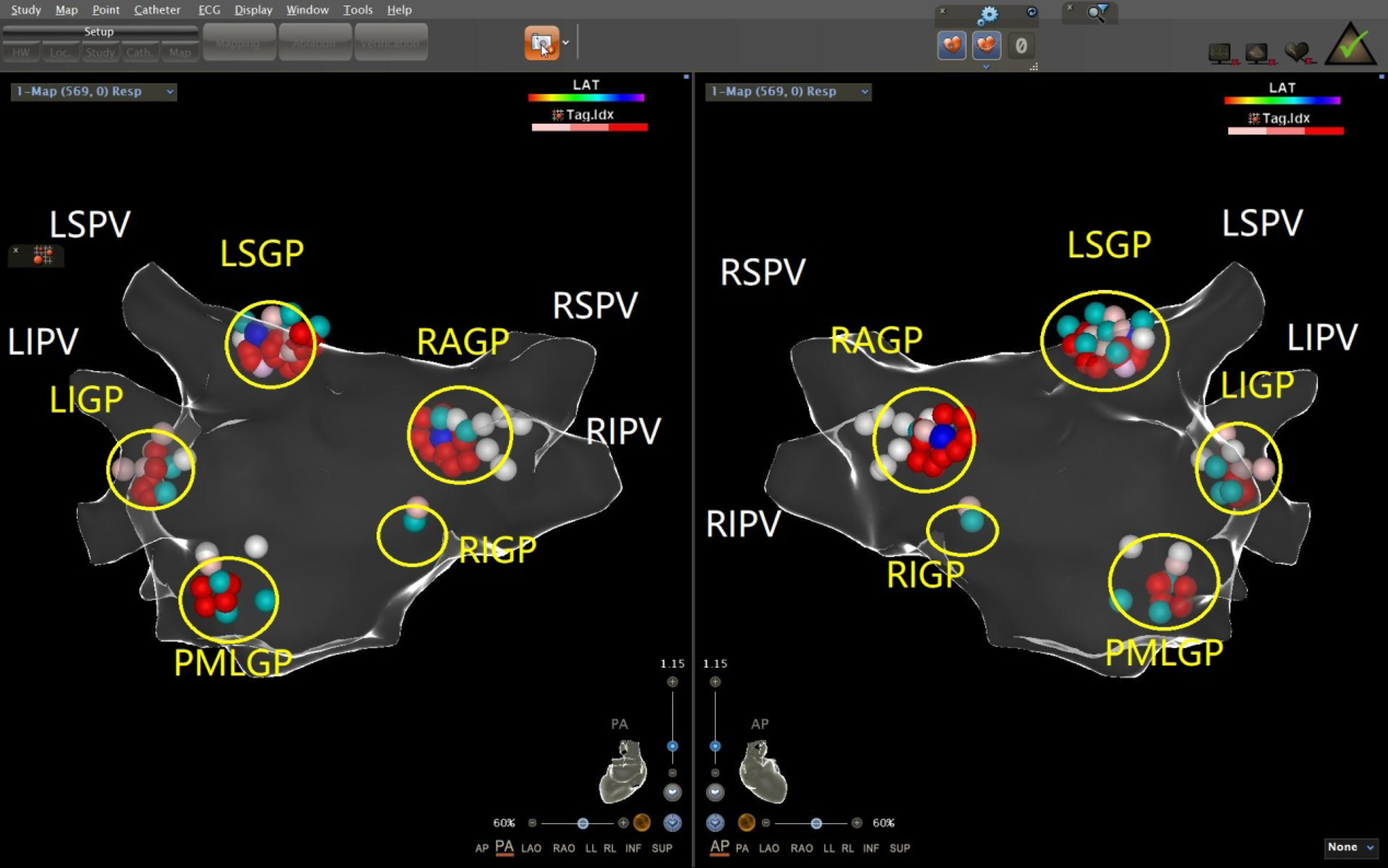
Schematic representation of the distribution of the autonomic ganglionated plexus in the left atrium of a patient with vasodepressant VVS

**Figure 3.**
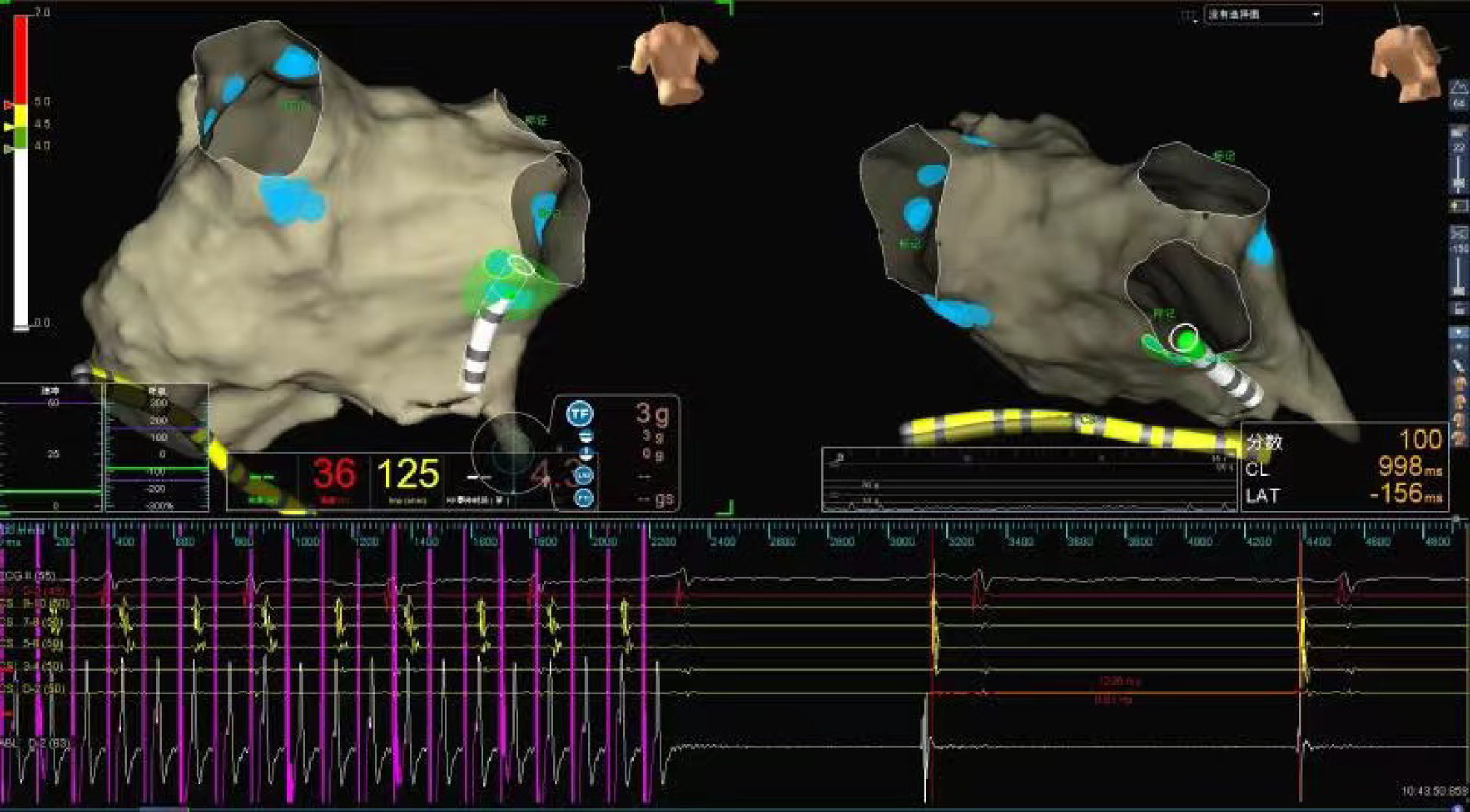
Schematic representation of cardiac ganglionated plexus ablation in a patient with VVS

Heart rate and electrophysiological parameters of GP ablation targets before and after ablation in patients with mixed and vasodepressor VVS are shown in Table 2 and Figure 4. In both mixed and vasodepressor VVS, vagal responses were most frequently observed at the LSGP and RAGP. Responses were observed at LSGP, LIGP, PMLGP, RAGP, and RIGP in 19 (100%), 17 (89.47%), 8 (42.11%), 16 (84.21%) and 18 (94.74%) patients, respectively. Among the 13 patients with vasodepressor VVS, vagal responses were observed at the LSGP, LIGP, PMLGP, RAGP, and RIGP in 12 (92.30%), 12 (92.30%), 5 (38.46%), 13 (100%), and 13 (92.30%), respectively. The number of ablation targets did not significantly differ between the vasodepressor and the mixed types of VVS. LSGP was the site where vagal responses were most commonly observed during HFS. No serious complications such as cardiac perforation or pericardial effusion occurred.

**Figure 4.**
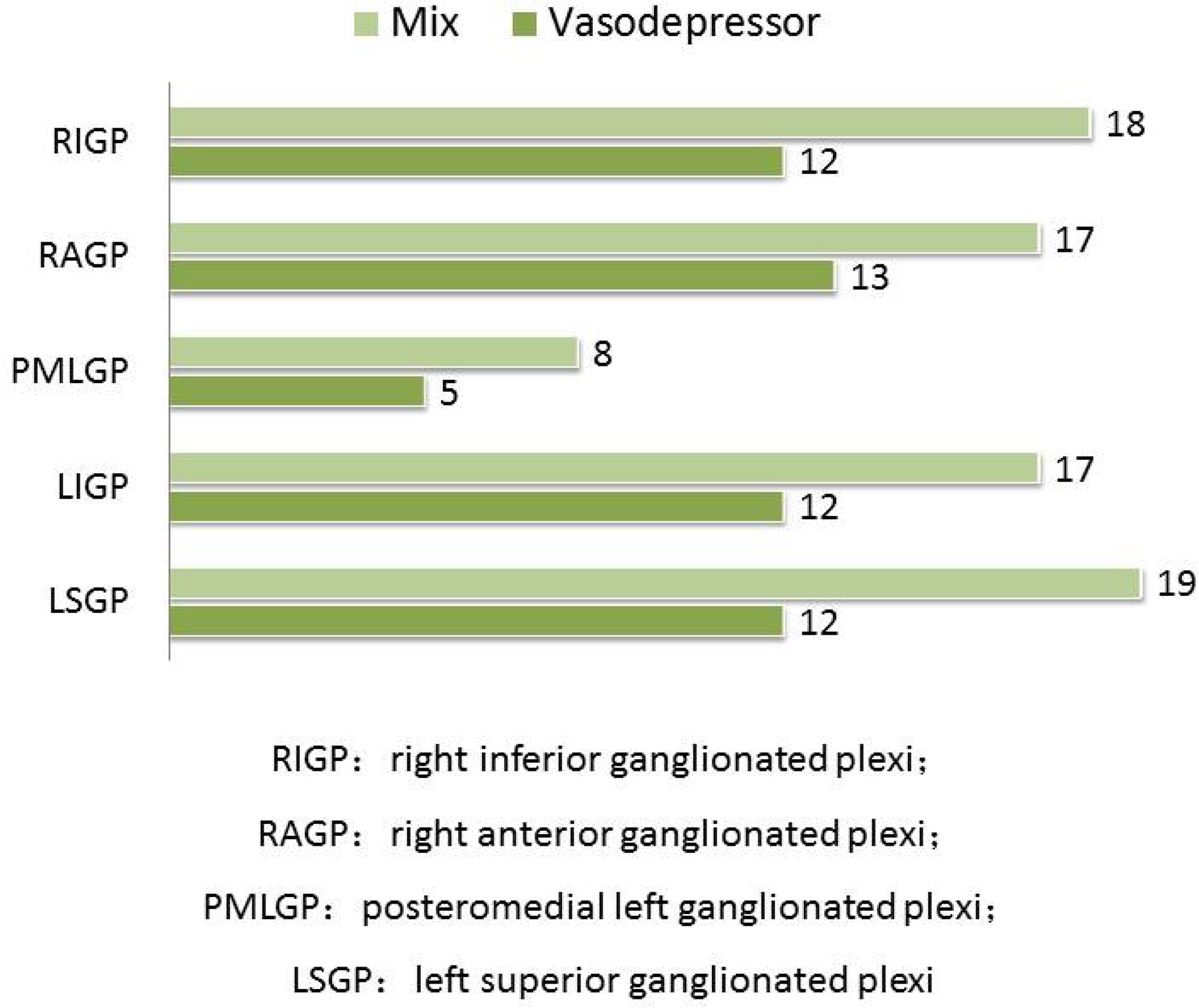
Comparison of the frequency of positive vagal reaction during GPs ablation

**Table 2.** Heart rate and electrophysiological parameters before and after ablation.

| Type |  | Before ablation | After ablation | t | P |
| --- | --- | --- | --- | --- | --- |
| Vasodepressor type | Heart rate | 67.80±12.22 | 84.17±15.91 | -4.317 | 0.002 |
|  | Venn's point of AV conduction (ms) | 446.25±115.73 | 350.00±75.19 | 4.934 | 0.001 |

In the vasodepressor VVS group, mean preoperative heart rate was 67.80 ± 12.22 beats/minute and the Wenckebach point of atrioventricular conduction was 446.25 ± 115.73 ms. After ablation, the mean preoperative heart rate was significantly higher (84.17 ± 15.91 beats/minute) and the Wenckebach point was significantly lower (350.00 ± 75.19 ms), as shown in Table 3.

**Table 3.** Heart rate and heart rate variability parameters on Holter monitoring before and after ablation.

| Type |  | Before ablation | 3 months after ablation | P |
| --- | --- | --- | --- | --- |
| Vasodepressor type | Max heart rate | 120.00±17.68 | 123.15±16.79 | <0.001 |
|  | Minimum heart rate | 49.23±8.69 | 58.31±11.40 | <0.001 |
|  | Mean heart rate | 69.15±9.33 | 76.54±9.52 | <0.001 |
|  | SDNN | 144.71±44.63 | 89.36±36.14 | <0.001 |
|  | SDANN | 127.47±44.21 | 81.75±30.43 | <0.001 |
|  | RMSSD | 41.11±16.56 | 24.61±15.87 | <0.001 |
|  | PNN50 | 10.65±8.55 | 5.52±6.55 | 0.025 |

SDNN: Standard deviation of the NN interval;
SDANN: Standard deviation of average NN interval;
RMSSD: Squares of differences between adjacent NN intervals;
PNN50: adjacent NN intervals that are greater than 50ms

### FOLLOW-UP

Mean follow-up was 11 ± 4.67 months. Syncope recurred in two of 13 patients with vasodepressor VVS after treatment, one at 3 months and the other at 8 months, one of them had prodromal symptoms after ablation, and the symptoms were relieved compared with those before ablation. The remaining patients did not experience any syncope or prodromal symptoms (Table 4). In patients with vasodepressor VVS, mean heart rate on 24-hour Holter monitoring increased from 67.80 ± 12.22 beats/minute to 84.17 ± 15.91 beats/minute after ablation. Among the HRV indexes, Standard deviation of the NN interval (SDNN), Standard deviation of average NN interval (SDANN), Standard deviation of the NN interval index (SDNNi), Squares of differences between adjacent NN intervals (rMSSD), and adjacent NN intervals that are greater than 50ms (pNN50) were significantly lower 3 months after ablation (P <0.001), as shown in Table 5. No serious adverse reactions such as cardiac tamponade, pulmonary embolism, or arrhythmia occurred after ablation. One patient developed sinus tachycardia 20 days after ablation, which was successfully treated with oral metoprolol.

**Table 4.** Follow-up results of syncope symptoms in two groups of patients with different types of VVS.

| Type | Postoperative recurrent syncope or presyncope | There was no recurrence of syncope after operation | P |
| --- | --- | --- | --- |
| Mixed type | 1 (%) | 18 (%) | 0.194 |
| Vasodepressor type | 3 (%) | 10 (%) |  |

**Table 5.** Heart rate and HRV time domain parameters of holter were recorded before and after ganglionated plexus ablation.

| Type |  | Before ablation | 3 months after ablation | P |
| --- | --- | --- | --- | --- |
| Mixed type | Max heart rate | 116.11±17.25 | 119.11±16.38 | <0.001 |
|  | Minimum heart rate | 52.67±8.57 | 66.28±11.49 | <0.001 |
|  | Mean heart rate | 71.61±9.43 | 83.94±9.65 | <0.001 |
|  | SDNN | 116.87±42.94 | 60.85±36.71 | <0.001 |
|  | SDANN | 102.87±42.73 | 64.23±30.91 | 0.001 |
|  | RMSSD | 35.55±16.81 | 16.07±16.04 | 0.001 |
|  | PNN50 | 8.12±8.64 | 1.10±6.63 | <0.001 |
| Vasodepressor type | Max heart rate | 120.00±17.68 | 123.15±16.79 | <0.001 |
|  | Minimum heart rate | 49.23±8.69 | 58.31±11.40 | <0.001 |
|  | Mean heart rate | 69.15±9.33 | 76.54±9.52 | <0.001 |
|  | SDNN | 144.71±44.63 | 89.36±36.14 | <0.001 |
|  | SDANN | 127.47±44.21 | 81.75±30.43 | <0.001 |
|  | RMSSD | 41.11±16.56 | 24.61±15.87 | <0.001 |
|  | PNN50 | 10.65±8.55 | 5.52±6.55 | 0.025 |

SDNN: Standard deviation of the NN interval;
SDANN: Standard deviation of average NN interval;
RMSSD: Squares of differences between adjacent NN intervals;
PNN50: adjacent NN intervals that are greater than 50ms

## DISCUSSION

This study suggests that CNA of the left atrial GPs is safe and effective in patients with VVS. Data were consistent across study patients, suggesting good reproducibility. All patients had symptoms of syncope before treatment and experienced a significant increase in minimum and mean heart rate and improvement in symptoms after CNA.

### Safety and effectiveness of CNA in treating VVS

Radiofrequency ablation of the vagal plexus has become a new strategy for treating VVS. Through the continuous research of various medical teams and advances in technique and technology, the procedure has shown promise in VVS patients. In this retrospective study, 13 patients with vasodepressor VVS were treated with radiofrequency ablation using HFS combined with anatomical localization. During the procedure, radiofrequency ablation was performed on five common vagal ganglia. Dynamic electrocardiography showed that HRV indexes were significantly lower at the 3-month follow-up than before the procedure. At the 12-month follow-up, only two patients had experienced recurrent syncope and one had experienced prodromal symptoms. The others reported no recurrence or physical injury. Our results show that radiofrequency ablation of the vagal plexus is effective and safe in patients with VVS. The autonomic nervous system affects the function of the cardiovascular system by regulating the balance between sympathetic and parasympathetic tone. VVS is caused by the imbalance of sympathetic and parasympathetic input and the negative effect of pathological vagal nerve tone on cardiac conduction and vascular tone. In two studies, Pachon et al. showed that permanent endocardial denervation can be achieved to treat VVS by catheter ablation of epicardial ganglia in the atrial wall from the endocardial surface[4, 11]. Clinical research has demonstrated that catheter ablation can significantly improve symptoms in patients with VVS[10].

In the atrium, parasympathetic nerves are more densely distributed than sympathetic nerves and mainly located throughout the subendocardium. The GPs in the left atrium are mainly located around the roots of the pulmonary veins. Sun[12] et al. performed GP ablation only in the left atrium, which had good long-term clinical results: postoperative head-up tilt testing changed from positive to negative in 81.5% of patients and 79.1% changed from mixed inhibition to negative; 11.9% changed from mixed inhibition to vascular inhibition, suggesting that catheter ablation is more effective for mixed inhibition of VVS and can effectively relieve the inhibition of heart rate. The vagal response frequency at LSGP and RIGP was significantly higher in patients with mixed and vasodepressor VVS during ablation, suggesting that these should be the main ablation targets, which is consistent with the results of our study.

It is also crucial to accurately determine the end point of radiofrequency ablation of the vagal plexus. In our study, HFS was used to directly stimulate the vagus nerve when determining the ablation end point. A negative vagal response proved that the ablation end point was reached.

### Effectiveness of CNA on different types of VVS

The recurrence of syncope in 35 patients with VVS after cardiac radiofrequency ablation was compared. We found no significant difference in long-term effectiveness between the mixed and vasodepressor types after ablation, indicating that cardiac GP radiofrequency ablation is equally suitable for both. We divided VVS patients into cardioinhibitory, vasodepressor, and mixed types according to head-up tilt testing results. The reason for the hemodynamic differences in VVS patients is still unclear, but may be related to different sensitivities of neuromodulation of the heart and blood vessels. Our study showed that during head-up tilt table testing, there was no difference in baseline blood pressure, heart rate, mean vascular resistance, or cardiac index between the mixed and vasodepressor types. However, when syncope occurred, mean vascular resistance in the vasodepressor type group significantly decreased and the heart rate and cardiac index significantly increased. In contrast, mean vascular resistance, heart rate, and cardiac index decreased in the mixed type group. The balance of sympathetic and vagal tone can be reflected by heart rate and vascular resistance is mainly controlled by sympathetic input[13]. Folino[14] et al. found in a study of the regulation of autonomic nervous activity in head-up tilt-induced syncope that HRV did not significantly differ between VVS patients and controls in the basal state[15]. Before syncope, sympathetic activity was increased in patients with cardioinhibitory and mixed types of VVS; however, heart rate decreased because of stronger vagal activity. Patients with vasodepressor type VVS have decreased vascular tone because of insufficient sympathetic activation. Radiofrequency ablation of the vagal plexus has a therapeutic effect on patients with VVS by inhibiting vagus nerve function. Therefore, it has a better effect on patients with cardioinhibitory and mixed VVS, which are caused by excessive activation of vagus nerve function. Xia[10] et al. found significant improvement in symptoms of cardiac depression but not vascular depression when the vagal reflex was repeatedly induced after ablation of the epicardial fat pad in dogs. Their analysis was driven by the fact that radiofrequency ablation of the cardiac GP significantly attenuated the cardiac vagal component but had no effect on the vasodepressor component of the Bezold-Jarisch reflex. Studies have suggested that although CNA can significantly improve the symptoms related to cardiac depression, the improvement of vagus depression symptoms has not reached the ideal effect. Therefore, VVS patients should be evaluated and classified to formulate an individualized treatment plan before CNA is performed[16]. These results suggest that radiofrequency GP ablation is more effective for cardioinhibitory and mixed VVS and less effective for vasodepressor VVS. This differs from our results, as we found that CNA has the same effectiveness in patients with vasodepressor VVS. There may have been differences in GP localization and ablation accuracy between different operators in our study. In addition, the sequence of ablation targets in our study was not necessarily the same between operators. Studies have shown that different sequences may affect efficacy of ablation. Finally, because the patients did not have an implantable electrocardiogram recorder, the cause of their syncope at the time of recurrence is not known. Future randomized large-scale trials are needed to study the effectiveness of vagal plexus radiofrequency ablation for treating the different types of VVS.

### Causes of syncope recurrence after CNA treatment of VVS

In this study, the LSGP was the most common ganglia with vagal responses. Previous studies have found that the left atrial GPs are distributed primarily around the pulmonary vein orifice, especially the junction between the left superior pulmonary vein and left atrium. In an autopsy study, Chevalier[17] et al. found that GPs in the left atrium were mainly distributed at the proximal end of the left and right pulmonary veins and the junction of the atrium; in addition, the left atrium was more highly innervated than the right[18]. Therefore, radiofrequency ablation of the GPs of the left atrium is usually used in clinical practice. However, right atrial ablation has also been reported. Armour et al. found that LSGP and RAGP had a significantly greater number of ganglia distributed than LIGP and RIGP[19]. Pachon et al. found that the LSGP is the GP with the highest distribution density in the left atrium and plays a key role in autonomic regulation of the atrioventricular node[20, 21]. Stimulation of the LSGP can cause negative time and conduction effects on the heart, which can be significantly reduced after LSGP ablation. In an ablation study of patients with atrial fibrillation, the vagal response was more common on the left atrial side of the pulmonary vein around the ostium, and the vast majority of the vagal responses occurred in the LSGP. The results of our study were similar. Ablation points during CNA are mainly concentrated in the GP on the atrial septal side, while the degree of ablation around the pulmonary vein ostium, especially the LSGP, was less or not ablated. This incomplete denervation may be the reason for syncope recurrence after VVS[13, 22]. In our study, three patients experienced recurrence of syncope or prodromal symptoms after ablation; however, the prodromal symptoms were improved. In some patients who underwent head-up tilt testing, the syncope induction time was longer than before, because vascular depression could not be completely eliminated by cardiac ablation; however, the late vascular depression was significantly delayed and weakened compared with before ablation. Because of interoperator differences, the degree of vagal plexus ablation and vagal denervation may have been different or incomplete. Several studies have shown that a similar situation may occur in patients who have undergone pacemaker implantation. In these patients, vascular depression-induced syncope may occur despite the prevention of cardiac arrest by the pacemaker[23]. Even if residual vasodepressor syncope is present, it can be cured by radiofrequency ablation of the cardiac GP to increase the threshold of symptoms.

### Limitations

This study has several limitations. First, it was retrospective in design so our results need to be confirmed in a large prospective randomized controlled trial. Second, implantable electrocardiogram recording was not used after treatment so the exact mechanism of syncope recurrence is not well understood.

## CONCLUSION

This study confirmed the effectiveness of radiofrequency GP ablation for preventing recurrence of VVS, providing a theoretical basis for individualized treatment of VVS patients. HRV indexes after ablation were significantly lower, indicating that CNA can achieve good results for vasodepressor type VVS. However, further large-scale prospective randomized controlled studies are needed.

## Data availability statement

The data that support the findings of this study are available on request from the corresponding author. The data are not publicly available due to privacy or ethical restrictions.

## Acknowledgment

We thank Liwen Bianji (Edanz) (https://www.liwenbianji.cn) for editing the English text of a draft of this manuscript.

